# A knowledge, attitude and practices study on malaria in Kandhamal district, Odisha

**DOI:** 10.1101/2023.07.11.23292508

**Authors:** Indumathi Mohan, Meenaketan Das, Meenu Mariya James, Rashi Dixit, Sujit Kumar Behera, Gunanidhi Dhangadamajhi, Prakash Babu Kodali, Karuppusamy Balasubramani, Praveen Balabaskaran Nina

**Affiliations:** Department of Epidemiology and Public Health, School of Life Sciences, Central University of Tamil Nadu, Tiruvarur, India-610005; Department of Biotechnology, Maharaja Sriram Chandra Bhanja Deo University, Baripada, Odisha, India- 757003; Department of Public Health and Community Medicine, Central University of Kerala, Kasaragod, Kerala, India- 671316; Department of Geography, School of Earth Sciences, Central University of Tamil Nadu, Tiruvarur, India-610005

**Author notes:** Correspondence: Dr. Praveen Balabaskaran Nina, Department of Public Health and Community Medicine, Central University of Kerala Kasaragod, Kerala, India, Email ID. Equal contribution (Indumathi Mohan, Meenaketan Das, Meenu Mariya James).

**Keywords:** Malaria, Kandhamal district, KAP study, Scheduled tribes and malaria, Odisha and malaria, GIS

## Abstract

Malaria is endemic in several tribal-dominated districts in India. Partnering and educating the tribal communities are key to malaria elimination efforts. A Knowledge, Attitude, and Practices (KAP) study was undertaken at Kandhamal, a tribal-dominated malaria endemic district in the state of Odisha, to assess the gaps in malaria awareness in this community for focused public health interventions. To assess the knowledge, attitudes, and practices on malaria in the tribal-dominated regions of Kandhamal district, Odisha. A descriptive KAP survey was carried out in the Kandhamal district at the household level. Three hundred households/respondents from 6 blocks distributed in 44 villages were selected through convenient sampling. Statistical analysis was carried out using SPSS (version 24), and ArcGIS software was used for GIS mapping. The respondents had good knowledge on the major malaria symptoms. Witchcraft, contaminated food/water, and contact with a malaria case were thought to transmit malaria by 5.3%, 14.3%, and 9.7% of respondents, respectively, and these clusters have been mapped by GIS. Logistic regression identified reduced level of education and open source of water supply to have a strong association with the misconceptions regarding malaria among the respondents. Knowledge and attitude regarding malaria were found to be associated with residence in a hilly area and an open source of water supply, respectively. Overall, the participants in Kandhamal had good KAP on malaria. The clusters with poor knowledge could be the target of focused public health interventions

## Introduction

Malaria, an acute febrile illness caused by the protozoa *Plasmodium*, is a vector-borne tropical disease. In 2020, 241 million malaria cases worldwide were reported, with 0.6 million deaths.[1] Two percent of the global malaria burden is shared by the WHO South East Asia region.[1] India contributes to 83% of the malaria burden in this region, and more than one-third of all cases are *P. vivax.*[1] India’s tropical climate and geographical topology favor perennial malaria transmission in many regions.

Historically, the state of Odisha, located on the east- coast of the Indian peninsula, is highly endemic to malaria. [2, 3] Odisha’s two geophysical locations, the northern plateau, and eastern ghats provide favorable environmental conditions for malaria transmission.[4] Odisha’s hot and humid climate, along with its forested regions[5], favors meso- to the hyper-endemic transmission of malaria.[6] In 2017, the Government of Odisha initiated the Durgama Anchalare Malaria Nirakarana (DAMaN) program to carry out a half-yearly mass survey in hard-to-reach malaria- endemic pockets of 23 districts. Even though the *P. falciparum* cases steeply decreased by >80% after the launch of DAMaN, several pockets in Odisha continue to have a significant malaria burden.[7] Furthermore, malaria elimination efforts in parts of Odisha are hindered by hard-to- access hilly and densely forested regions mainly inhabited by indigenous tribal groups.[4, 8] In addition to being a malaria-endemic state, Odisha is one of the top ten tribal-inhabited states.[9] The 2011 census reports 22.8% of the total population of Odisha is comprised of tribals; tribal groups constitute 62% of the entire rural population of Odisha.[10] Past studies have well documented the disproportionate burden of diseases in the Indian tribals,[11] including malaria.[9]

India has set an ambitious goal of malaria elimination by 2030. Good behavior, attitude, perceptions, and knowledge regarding malaria, especially in vulnerable tribal communities, are essential for effectively implementing malaria elimination strategies. Studies on Knowledge Attitude and Practice (KAP) will help understand the misconceptions and disbeliefs that influence the knowledge and attitude of the population in malaria-endemic tribal regions. Kandhamal is one of the highly malarious districts in Odisha.[3] Scheduled Tribes (ST) make up 53.5% of the population in the Kandhamal district.[12] There is no available literature on malaria KAP studies in this district. Here, we detail the KAP study carried out to understand the beliefs, perceptions, attitudes, knowledge, and practices concerning malaria in this district; the study outcome may supplement the malaria elimination efforts of the state Govt.

## Methods

### 1. Study site

Kandhamal district in Odisha state lies between 19 degrees 34’ to 20 degrees 36’ north latitude and 83 degrees 34’ to 84 degrees 34’ east longitude. The district covers an area of 7654 sq km and is divided into 12 blocks[12] [Fig. 1(A)]. The temperature ranges from 45.5 degrees C to 2.0 degrees C, with an average annual rainfall of 1500 mm.[12] The entire district lies in an undulated terrain with hilly ranges and narrow valley tracts. Almost 2/3 of the district is covered with dense forests. The topography of the district restricts the land use, economic activities, and development of the district.

**Figure 1:**
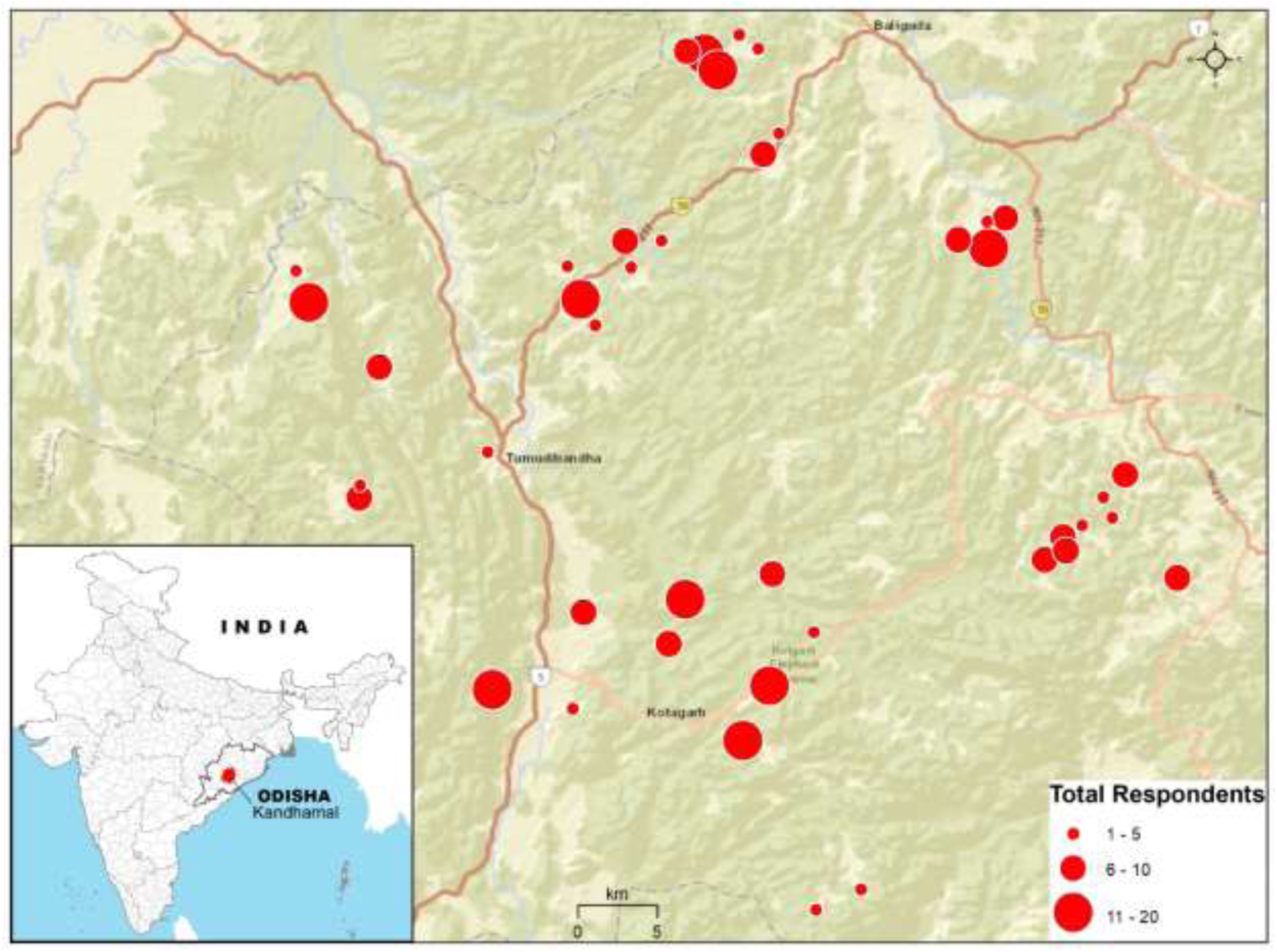
Site Map: Location of study area and distribution of surveyed villages and total respondents from each village. The size of the circles is proportional to the total number of respondents. The background of the map is obtained from ArcGIS imagery services for representing topography, land use/ land cover and accessibility of the study area.

### 2. Study design and population

The study design is a descriptive cross-sectional study. Out of 12 blocks in Kandhamal district, six blocks (Balliguda, Nuagaon, Daringbadi, Tumudibandh, Bergarh, and Kotagarh) were conveniently selected for this KAP study [Fig. 1]. In these 12 blocks, based on the population size, 44 villages have been selected. Furthermore, 300 households from these 44 villages were conveniently selected. One household was taken as one sampling unit, and one representative from each household was considered a respondent.

### 3. Data Collection

A structured questionnaire was prepared and used for data collection from the selected households by a research assistant. The participants were either the head of the family or the primary decision-maker of the household. If the head of the household was absent, adults>18 years of age selected by the household members were asked to participate in the survey. The questionnaire contained 51 questions (close-ended), divided into two main sections: Section A - participant’s details and section B - KAP of malaria. Questions in section A were mainly about the participant’s basic details, demographic and socio-demographic indicators. Section B comprised KAP questions on symptoms and causes, transmission, treatment and access to healthcare, prevention and control practices, vector control and management, and Information, Education, and Communications (IEC) activities. Originally drafted in English, the questionnaire was administered after translation to Odia. The duration of data collection was disrupted due to the COVID-19 pandemic; hence the data collection was done for an extended period between April- August 2021. A full verbal explanation was given to the participants, and written consent was acquired before participation.

### 4. Statistical analysis

Data from the questionnaire was manually entered into Microsoft Excel. All the data were statistically analyzed using SPSS version 20.0. Frequency statistics were performed for descriptive analysis of data. Three dependent variables were developed to perform logistic regression, as done in earlier KAP studies.[13–17] (i) Knowledge Score (*K*) [*K* score was created using three variables (questions) that study knowledge. They are: 1. To correctly know what transmits malaria 2. To correctly know what time the malaria mosquito bites, and 3. To correctly affirm if malaria is treatable]; (ii) Attitude Score (*A*) [*A* score was created using three variables (questions) that study attitude. They are: 1. To identify how soon they will seek treatment, 2. What is the best treatment for malaria according to them, 3. Is the fight against malaria elimination a collective responsibility of the community?]; (iii) Misconception scoring was created using five variables. They are - to identify (1) Witchcraft, (2) consuming contaminated food, (3) contact with a malaria case, and (4) dirty drinking water as the transmission mode of malaria, (5) seeking treatment from a traditional healer. For both *K* and *A* scores, a score of =3 was deemed good; a score <3 was deemed poor. For the misconception score, a score of ≥1 was deemed as having misconceptions; a score of 0 was deemed as having no misconceptions [refer to supplementary table 1 for the scoring scheme]. Logistic regression analysis was carried out to identify potential predictors associated with participant’s knowledge, attitude, and misconceptions. An alpha level <0.05 was considered statistically significant throughout the analysis.

### 5. Mapping of Clusters

The locations of surveyed were obtained using GNSS (Global Navigation Satellite System) receiver, and the coordinates were transferred into the GIS (Geographical Information System) database using ArcGIS software for mapping. The number of respondents was aggregated village- wise and prepared as a graduated symbol map to understand the respondents’ spatial distribution (Figure 1). The parameters used to understand the level of misconception about malaria, such as dirty drinking water, contaminated food, witchcraft, contact /touch, and traditional healer, were individually mapped using proportional symbols (Figure 3). The number of respondents of each parameter is compared with the total number of respondents using an overlay technique, and clusters of misconception are identified.

## Results

### 1. Socio-demographic characteristics

Table 1 shows the demographic and socio-economic characteristics of the study population. Of the 300 respondents, the majority (79%) were males. 67.3% (202) of the respondents were observed to have no schooling or only received primary education, and 32.7% (98) had secondary education and above. Furthermore, the highest number (80%) of respondents were employed in income-generating activities, and 20% were in the non-earning group. Almost all the respondents (99%) reside in a kutcha house, and the majority (80.3%) live in hilly areas. The major water source is piped supply for 79.7% of the households.

**Table 1:** Demographic and socio-economic characteristics of study participants in selected households of Kandhamal dist. Odisha, 2021.

| Indicators |  | N= 300 | % |
| --- | --- | --- | --- |
| **Gender** | Male | 237 | 79 |
|  | Female | 63 | 21 |
| **Age** | <45 years | 193 | 64.03 |
|  | >45 years | 107 | 35.7 |
| **Education** | No schooling/ Primary | 202 | 67.3 |
|  | Secondary education and above | 98 | 32.7 |
| **Occupation** | Earning | 240 | 80 |
|  | Non-earning | 60 | 20 |
| **Religion** | Hindu | 60 | 80 |
|  | Christian | 240 | 20 |
| **Annual Income** | <Rs.1,00,000 | 397 | 99 |
|  | >Rs.1,00,000 | 3 | 1 |
| **Marital Status** | Married | 298 | 99.3 |
|  | Unmarried | 2 | 0.7 |
| **Family Structure** | Nuclear | 256 | 85.3 |
|  | Joint | 44 | 14.3 |
| **Kind of House** | Kutcha | 297 | 99 |
|  | Pucca | 3 | 1 |
| **Geographic location of the house** | Forest | 35 | 11.7 |
|  | Hilly area | 241 | 80.3 |
|  | Plains | 24 | 8 |
| **Type of water supply at home** | Open source | 61 | 20.3 |
|  | Piped | 239 | 79.7 |
| **Whether practice Animal husbandry or not** | Yes & rear bovines | 294 | 98 |
|  | No | 6 | 2 |
| **Resting place of live stock** | Inside the house | 32 | 10.7 |
|  | Outside the house | 241 | 80.3 |

### 2. KAP survey: Symptoms and causes

The questionnaire and the replies to queries on malaria KAP is given in Table 2. Many of the study participants were aware of the major symptoms of malaria. All of them correctly identified fever as a symptom, and more than 80% correctly identified chills as a major symptom of malaria. Other symptoms of malaria, such as vomiting, lack of appetite, and headache, were identified by 15.3%, 12%, and 16% of the respondents, respectively [Fig. 2 (C)]. Almost all the respondents correctly associated mosquitoes with malaria. Among the respondents, self-reported malaria prevalence in the past one year was 16.6%.

**Figure 2:**
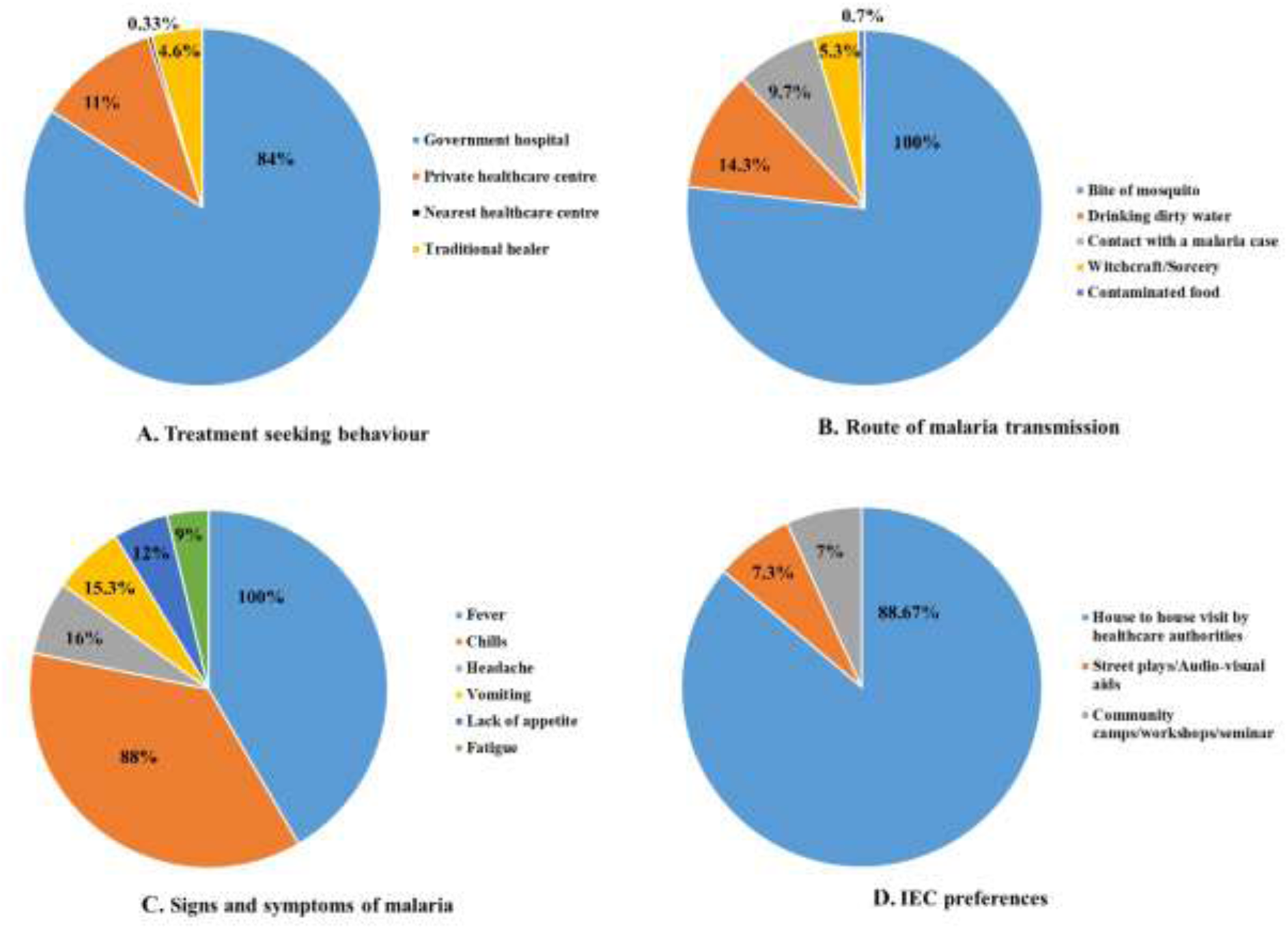
Pie charts depicting KAP concerning malaria reported by the participants in Kandhamal dist., Odisha. Figure 2 (A), 2 (B), 2 (C) and 2 (D) represent treatment-seeking behavior, route of malaria transmission, signs and symptoms of malaria, and IEC preferences of the participants, respectively. Sample size N=300. Frequency is expressed in absolute percentage (%). In Figure 2 (B) and 2 (C), frequency percentage might not add up to 100 because of multiple responses.

**Table 2:**
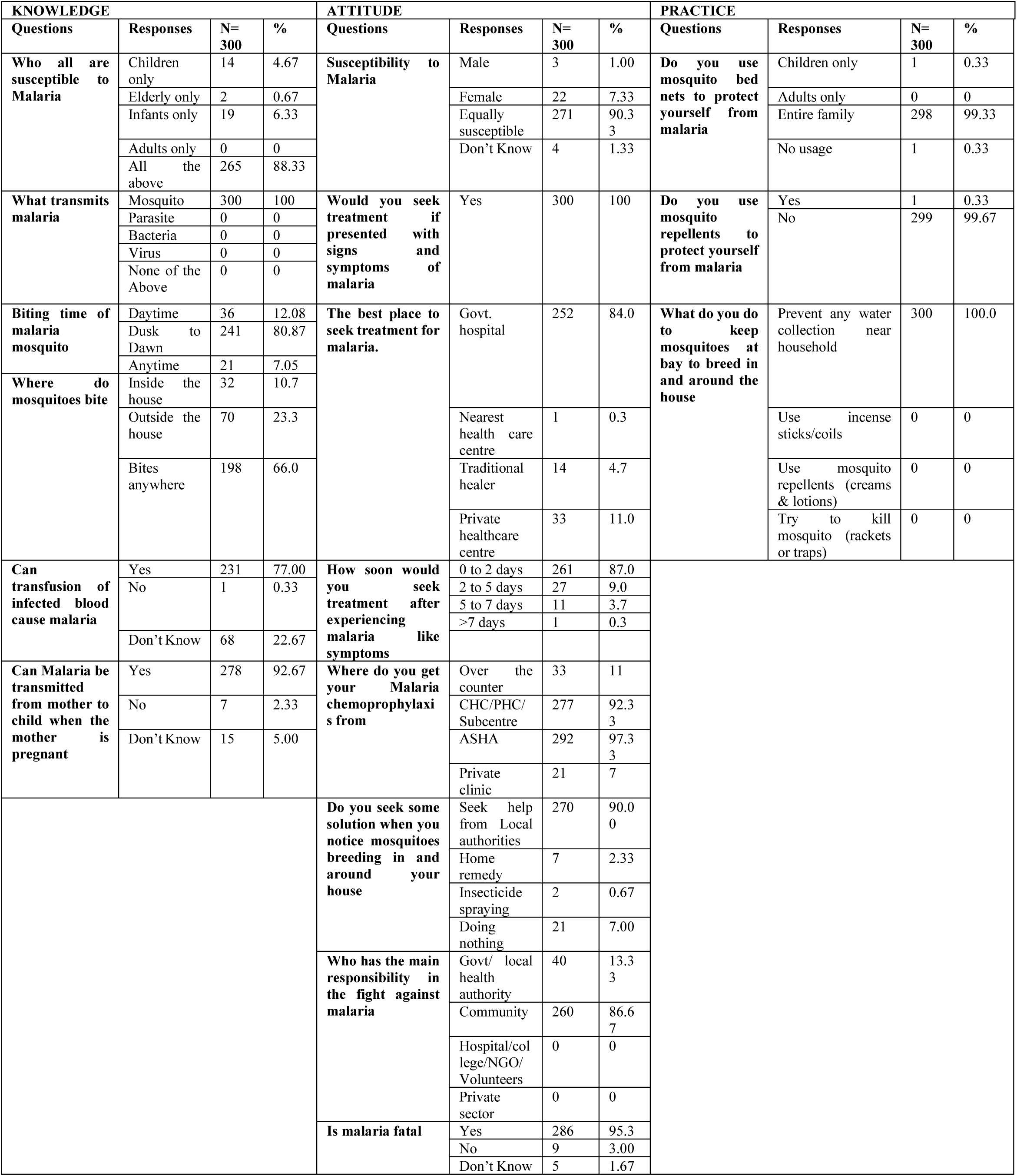
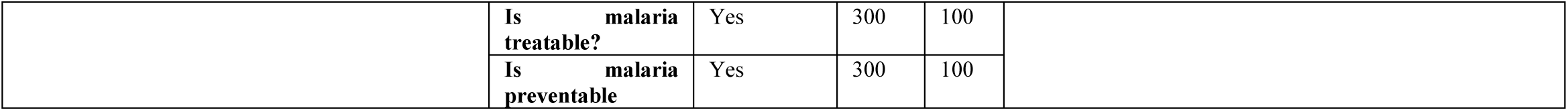
Malaria-related knowledge, attitude, and practices among the tribal in rural areas of Kandhamal district, Odisha, 2021.

### 3. KAP survey: Treatment and access to healthcare

All participants said they would seek treatment if they had symptoms of malaria. From the onset of symptoms, the majority (87%) said they would seek treatment within 0-2 days of being ill. The majority of the participants preferred a govt. hospital (84%), over a private facility (11%), for treatment [Fig 2(A)].

### 4. KAP survey: Information, education, and communication

House visits, street play/audio-visual aids, and community health camps were the preferred mode of IEC by 85.67%, 7.33%, and 7% of the respondents, respectively [Fig 2 (D)].

### 5. Misconceptions regarding malaria transmission

The majority of the study participants (84.3%) correctly linked the high occurrence of malaria to the monsoon season. However, some respondents had misconceptions regarding malaria transmission. Witchcraft, consuming contaminated food/water, and contact with a malaria case were thought to transmit malaria by 5.3%, 14.3%, and 9.7% of respondents, respectively [Fig. 2 (B)]. Respondents who withheld such misconceptions were scattered across the study site [Fig. 3]. However, the total number of such respondents is comparatively higher in the southern villages, and most of these villages are away from the major roads. Logistic regression was performed to estimate the relationship between the misconception among the respondents and their socio- demographic characteristics. The level of education was observed to have a strong association with the misconceptions regarding malaria among the respondents (Table 3). Compared to respondents with a secondary education level and above, respondents with no schooling or only primary education have higher odds of misconceptions regarding malaria (OR=1.9; 95% CI=1.02-3.5; P=0.043). Open source of water supply was also significantly associated with malaria misconceptions. Compared to respondents with piped water supply, respondents collecting water from an open source had higher odds of misconceptions regarding malaria (OR=2.18; 95% CI= 1.19-3.98; P=0.011) (Table 3).

**Figure 3:**
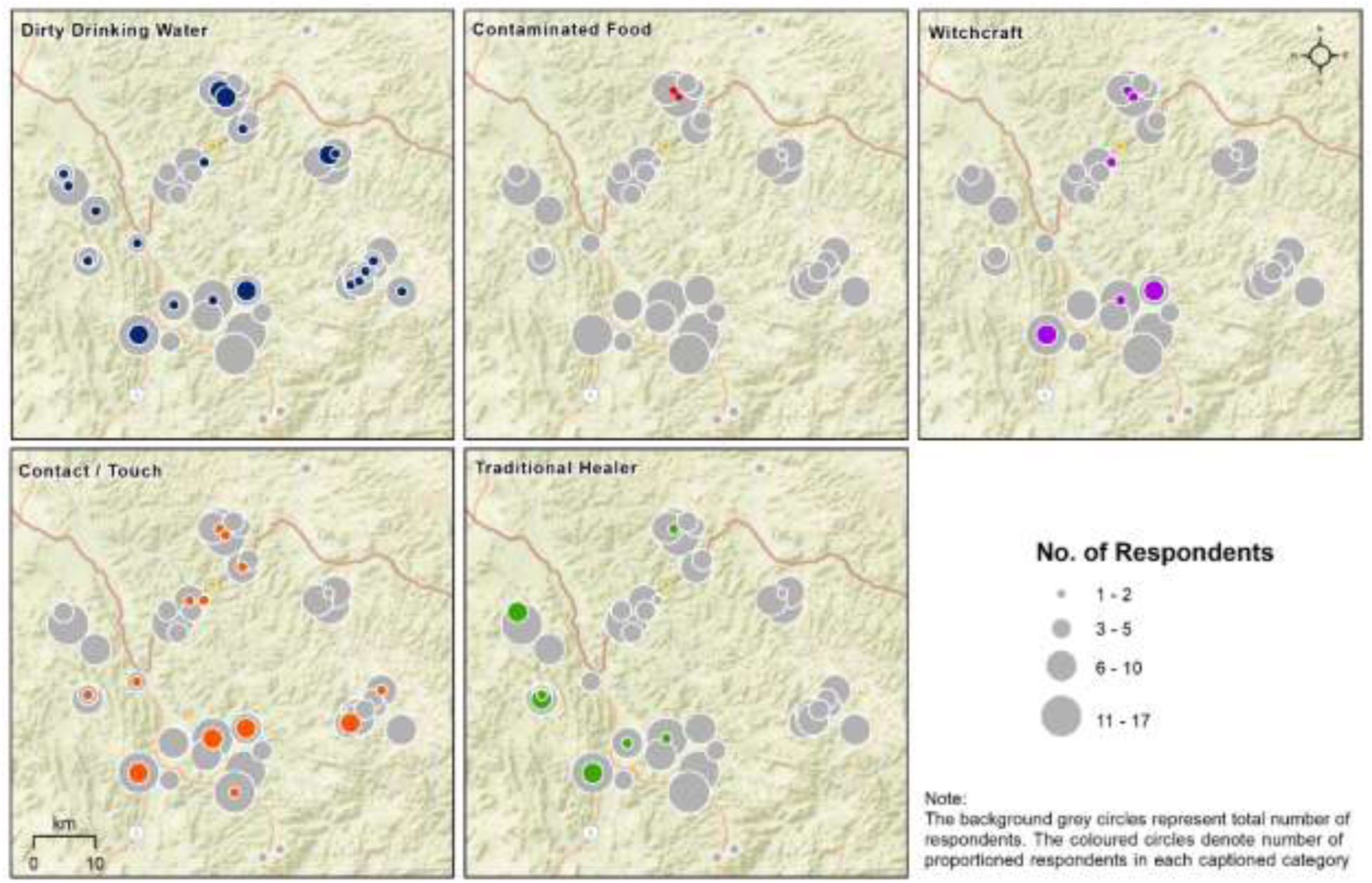
Misconception Map: Spatial distribution and clusters of misconception parameters. The size of the circles is proportional to the respondents of each category and overlaid on the total number of respondents (grey circles) for visual comparisons. The background of the map is obtained from ArcGIS imagery services for representing topography, land use/ land cover and accessibility of the study area. The major roads are represented with dark and thicker lines.

**Table 3:** Odds ratios of misconceptions, good knowledge and good attitude regarding malaria with socio-demographic variables.

|  | Misconception |  | Good Knowledge |  | Good Attitude |  |
| --- | --- | --- | --- | --- | --- | --- |
| Variables | OR (95% CI) | P-value | OR (95% CI) | P-value | OR (95% CI) | P-value |
| **Age** |  |  |  |  |  |  |
| <45 years | 1.15 (0.66, 2.0) | 0.62 | 0.7 (0.36, 1.3) | 0.28 | 0.79 (0.4, 1.4) | 0.47 |
| >45 years | Reference |  | Reference |  | Reference |  |
| **Gender** |  |  |  |  |  |  |
| Male | 0.38 (0.05, 2.7) | 0.34 | 1.5 (0.74, 3.14) | 0.24 | 1.2 (0.6, 2.5) | 0.55 |
| Female | Reference |  | Reference |  | Reference |  |
| **Level of education** |  |  |  |  |  |  |
| No schooling/Primary education | 1.9 (1.02, 3.5) | **0.043\*** | Reference |  | Reference |  |
| Secondary education and above | Reference |  | 1.5 (0.76, 3.3) | 0.21 | 0.57 (0.3, 1.07) | 0.08 |
| **Occupation** |  |  |  |  |  |  |
| Earning | 2.52 (0.32, 19.3) | 0.37 |  |  |  |  |
| Non-earning | Reference |  |  |  |  |  |
| **Type of water supply** |  |  |  |  |  |  |
| Open source | 2.18 (1.19, 3.98) | **0.011\*** | 0.61 (0.23, 1.54) | 0.29 | Reference |  |
| Piped | Reference |  | Reference |  | 4.09 (1.79, 9.3) | **0.001\*** |
| **Had history of malaria within past one year** |  |  |  |  |  |  |
| Yes | 1.32 (0.67, 2.61) | 0.41 |  |  | 1.19 (0.54, 2.65) | 0.65 |
| No | Reference |  |  |  | Reference |  |
| **Geographical characteristics of the location** |  |  |  |  |  |  |
| Forest canopy |  |  | 2.7 (0.67, 10.9) | 0.15 | 2.0 (0.43, 10.0) | 0.93 |
| Hilly area |  |  | 7.4 (2.9, 18.5) | **0.00\*** | 0.95 (0.30, 2.98) | 0.35 |
| Plains |  |  | Reference |  | Reference |  |

\*Significant at p<0.05

### 6. Socio-economic and socio-demographic determinants of KAP

Logistic regression analysis showed a strong association between the geographical characteristics of the respondents’ location and knowledge of malaria (Table 3). Compared to the respondents living in plain areas, hilly area residing respondents were observed to have higher odds of good knowledge (OR=7.4; 95% CI= 2.9-18.5; P=0.00). A significant association was also observed between attitude toward malaria and the water source. Respondents who had a piped supply of water had higher odds of having a good attitude on malaria when compared to respondents collecting water from an open source (OR=-4.09; 95% CI=1.79-9.3; P=0.001) (Table 3).

## Discussion

India is ramping up its efforts to eliminate malaria by 2030 through parasite and vector control strategies. For successfully implementing the strategies, it is imperative to actively educate and engage communities in the malaria-endemic pockets. A key step in this process is to assess the knowledge, attitude, and practices of malaria at the community level, especially where malaria is endemic. There are very few KAP studies on malaria in India, especially in the malaria endemic pockets. The current study carried out in the malaria-endemic, tribal-dominated Kandhamal district is one such effort to understand the status of malaria knowledge in the community.

Overall, the survey shows the households in Kandhamal have excellent knowledge of various aspects of malaria. Mosquito bite as a route of malaria transmission was correctly identified by the majority of the participants from this study, and our findings are in line with past KAP studies.[14, 18–22] An earlier community study carried out in 2006 in Boudh district in Odisha reported lower awareness on malaria among women and ST groups.[18] This positive shift in awareness and knowledge in Odisha can be attributed to the malaria control efforts by the Odisha government through the DAMaN programme started in 2017 as a supplementary initiative to strengthen malaria activities.[4] The three important pillars of malaria control in DAMaN are (i) Mass Screening and Treatment (MSAT), (ii) mass distribution and promotion of LLIN as a vector control strategy, and (iii) community mobilization through Behavioural Change Communication (BCC) in hard-to-reach remote villages.[4] The positive impacts of the second and third approaches are reflected in the results of this study, as evident by the good malaria knowledge, attitude, and practices (widespread use of bed nets).

Level of education was observed to be significantly associated with misconceptions about malaria in the present study. Deep cultural attachments and lack of adequate information from health workers, especially in tribal areas, usually cause such myths and misconceptions. In a descriptive cross-sectional survey conducted in rural Southwest Nigeria, the respondents’ low level of education was observed to be contributing to the misconceptions about malaria treatment.[23] Similarly, a study conducted in Northern Uganda also reported a significant association between the level of education and misconceptions regarding malaria.[24]

The respondents’ attitude towards malaria is positive, and the majority believe malaria can be prevented and controlled (Table 3). Studies conducted in northern India have also reported similar good attitudes among their study population.[14, 19] As reported in other studies from India, [14, 19, 25] the majority of the respondents preferred to get treated in a government hospital and will seek medical help within 0-2 days of a febrile episode. However, a small fraction (4.6%) of the respondents preferred to get treated by a traditional healer. Such health-seeking behaviour is not uncommon among tribal populations. A KAP study conducted in the tribal belt of Odisha (Koraput-Malkangiri) has reported the tribal’s preference to seek treatment from *Disharis* (traditional healers).[26] In addition to Accredited Social Health Activists (ASHA) and Village Health Nurses (VHN), IEC activities can also be implemented by partnering with and educating the village traditional healers[27] so that malaria intervention approaches can reach every household in the community.

Insecticide-treated bed nets are integral to preventing and controlling malaria, especially in hard- to-reach pockets in tribal regions, where hard-to-access terrains greatly impede access to healthcare and diagnostic facilities.[27] Here, the community was aware of the importance of the best practices for malaria control, and all the households were using bed nets. The overwhelming transformation to best malaria practices could be attributed to the DAMaN program. Malaria cases and Annual Parasite Index (API) steeply decreased in high-burden districts in Meghalaya after the introduction of LLINs in 2016.[28] Concerning malaria IEC preferences, the majority (88.6%) of the participants preferred house visits by local health authorities. Again, the heavy inclination towards house visits could be attributed to the DAMaN initiative, where a mass screening strategy is used to treat malaria cases in the remote pockets of the 23 malaria-endemic districts across Odisha.[4]

Culturally rooted misconceptions persisting in tribal populations are important sociological and geographical barriers to malaria control.[27] Nearly 10% of the respondents believed malaria could spread through touch/contact with a malaria patient. Also, ∼5% believed witchcraft and sorcery could spread malaria. Studies in the past have also reported similar misconceptions among their respondents.[26, 29–31] These misconceptions must be cleared for successful malaria elimination. The GIS approach used in this study can identify clusters where respondents with poor knowledge/misconceptions are dominant, and these clusters could be the focus of targeted interventions. Specific interventions will lead to greater acceptability among the tribes and greatly aid malaria elimination.

The study has important limitations. A pilot study before the survey could have greatly helped refine the questions. For example, the question on income had only two options: above or below $1300 per annum. Almost all the respondents were below the $1300 threshold. Ideally, a pilot study would have helped us stratify the income. Furthermore, the large set of questions (∼50) and the extended interview time (∼45 minutes per respondent) might have led to this study’s large dropouts.

Overall, the tribal-dominated study population in Kandhamal district, Odisha, possesses sound knowledge and attitude, and follows appropriate preventive practices against malaria. The GIS- based mapping could be useful for targeted public health intervention.

## Data Availability

All data produced in the present study are available upon reasonable request to the authors

## List of abbreviations

KAP: Knowledge Attitude and Practice
DAMaN: Durgama Anchalare Malaria Nirakarana
LLINS: Long-Lasting Insecticidal Nets
API: Annual Parasite Index
ST: Scheduled Tribes
MSAT: Mass Screening and Treatment
BCC: Behavioural Change Communication
IEC: Information, Education and Communication
ASHA: Accredited Social Health Activist
VHN: Village Health Nurse

## Acknowledgements

None

## Author Contributions

IM, MMJ, RD, and PBK analyzed and interpreted the data. RD wrote the first draft of the manuscript. MD carried out the survey. PBN conceptualized the study and wrote the manuscript. SKB and KB helped in interpreting the data. IM, MMJ, RD, MD, SKB, GD, PBK, BK and PBN contributed to the literature search, review and editing of the manuscript.

## Ethical Statement

The study was approved by the Human Institutional Ethical Review Board (IHERB) of the Central University of Tamil Nadu. Verbal consent was obtained from all the respondents who volunteered for the study.

