## Supplementary Table 1 for "A knowledge, attitude and practices study on malaria in Kandhamal district, Odisha"

**Supplementary Table 1:** Scoring scheme used to develop *K* score and *A* score.

<table>
<tr><th colspan="2">Variables used for knowledge scoring (K-score)</th><th>Correct knowledge<br>N (%)</th><th>Incorrect knowledge<br>N (%)</th><th>Good K-score<br>N (%)</th><th>Poor K-score<br>N (%)</th><th>Total<br>(N)</th></tr>
<tr><td>1.</td><td>Correctly knowing what transmits malaria</td><td>300 (100)</td><td>0</td><td rowspan="3">238 (79.3)</td><td rowspan="3">62 (20.7)</td><td rowspan="3">300</td></tr>
<tr><td>2.</td><td>Correctly knowing the biting time of mosquito that transmits malaria</td><td>238 (79.3)</td><td>62 (20.7)</td></tr>
<tr><td>3.</td><td>Correctly affirming that malaria is treatable</td><td>300 (100)</td><td>0</td></tr>
<tr><td colspan="7"></td></tr>
<tr><td colspan="2">Variables used for attitude scoring (A-Score)</td><td>Correct attitude<br>N (%)</td><td>Incorrect attitude<br>N (%)</td><td>Good A-score<br>N (%)</td><td>Poor A-score<br>N (%)</td><td>Total<br>(N)</td></tr>
<tr><td>1.</td><td>How soon the participants seek treatment?</td><td>261 (87)</td><td>39 (13)</td><td>238 (79.3)</td><td>62 (20.67)</td><td></td></tr>
</table>

| | | | | | | |
|---|---|---|---|---|---|---|
| 2. | The best treatment available for malaria according to the participants. | 299 (99.7) | 1 (0.3) | | | 300 |
| 3. | Susceptibility to malaria (sex-wise) | 271 (90.3) | 19 (9.6) | | | |
| **Variables used for misconception scoring** | | **Misconception (Yes)** **N (%)** | **Misconception (No)** **N (%)** | **Misconception (Yes)** **N (%)** | **Misconception (No)** **N (%)** | **Total** **(N)** |
| 1. | Witchcraft | 16 (5.3) | 284 (94.7) | 81 (27) | 219 (73) | 300 |
| 2. | Contaminated food | 2 (0.7) | 298 (99.3) | | | |
| 3. | Contact with a malaria case | 29 (9.7) | 271 (90.3) | | | |
| 4. | Dirty drinking water | 43 (14.3) | 257 (85.7) | | | |
| 5. | Traditional healer | 14 (4.7) | 286 (95.3) | | | |

2
